# Shared and distinct effects of cognitive performance and mood on cognition-dependent instrumental activities of daily living in Parkinson’s disease

**DOI:** 10.64898/2026.10.01.26364514

**Authors:** Jun Yu Hu, Sophie Sun, Madeleine Sharp

**Author notes:** Corresponding Author: Madeleine Sharp, 3801 rue University, Montreal, QC, H3A 2B4.

## Abstract

**Background:** Cognitive impairment in Parkinson’s disease (PD) significantly affects patients’ ability to perform instrumental activities of daily living (IADLs). However, neuropsychiatric symptoms, which are highly prevalent in PD, may also contribute to functional impairments. In this study, we investigated the independent contributions of cognitive and neuropsychiatric symptoms to functional impairments and tested whether mood mediates cognition–function associations.

**Methods:** We examined 104 patients from the Quebec Parkinson Network and replicated analyses in a sample of 144 patients from an independent online cohort. Participants completed the Penn Parkinson’s Daily Activities Questionnaire (PDAQ), a validated patient-reported measure designed to capture cognition-dependent IADLs, standard cognitive tests (executive function, attention, working memory), and neuropsychiatric questionnaires (apathy, depression, impulsivity, and anhedonia). Regression models tested the independent and combined effects of cognition and neuropsychiatric symptoms on PDAQ; mediation analyses explored mechanistic pathways.

**Results:** Both cognitive performance and neuropsychiatric symptoms were independently associated with PDAQ scores and the proportion of variance explained by neuropsychiatric symptoms was comparable to that explained by cognitive performance. Furthermore, depression and impulsivity partially confounded the relationship between cognitive performance and PDAQ and impulsivity partially mediated this relationship. The independent associations between neuropsychiatric symptoms and PDAQ were replicated in the replication sample.

**Conclusion:** These results indicate that impairments in cognitive IADLs reflect contributions of both cognition and neuropsychiatric changes and suggest that trials using measures of IADLs as an outcome measure should also carefully assess and control for neuropsychiatric symptoms.

**Plain language summary:** People with Parkinson’s disease often experience difficulties with everyday activities that rely on cognitive skills, such as organizing tasks, following instructions, or managing finances. These difficulties are usually linked to cognitive problems, but Parkinson’s disease is also commonly associated with mood and behavioral symptoms, including depression, apathy, and impulsivity. It is not well understood how much these symptoms contribute to problems with daily functioning compared to that of cognitive functions.

In this study, we examined the relationship between cognitive performance, mood symptoms, and daily functioning in two groups of people with Parkinson’s disease. Participants completed cognitive tests, questionnaires assessing mood and behavior, and the Penn Parkinson’s Daily Activities Questionnaire, which was designed to measure daily activities that depend on cognitive abilities. We found that both cognitive difficulties and mood symptoms were independently associated with worse daily functioning. Mood symptoms explained a similar amount of difficulty in daily activities as cognitive test performance. Depression and impulsivity partly confounded the relationship between cognitive impairment and daily functioning, and impulsivity also appeared to be one pathway through which cognitive difficulties affected everyday activities. Most findings were replicated in an independent group of participants.

These results indicate that difficulties in cognition-dependent daily activities in Parkinson’s disease reflect contributions from both cognitive impairment and mood or behavioral changes. Studies and clinical trials that use daily functioning as an outcome measure should therefore assess and account for mood symptoms to better understand whether changes in daily functioning are driven by cognitive improvement, changes in mood, or both.

## Introduction

Cognitive impairment in Parkinson’s disease (PD) is a major concern for patients and caregivers^1^. It greatly hinders patients’ ability to perform instrumental activities of daily living (IADL, e.g. navigating with a map, reading and following complex instructions, learning to use new gadgets or machines around the house)^2–5^, which are distinct from basic activities (e.g., bathing, dressing, and walking)^6^. The Penn Parkinson’s Daily Activities Questionnaire (PDAQ) is a well-established, reliable, and validated measure specifically designed to isolate and measure cognition-dependent IADLs in individuals with PD, making it a promising outcome measure for clinical trials targeting cognitive impairments in PD^7^. However, mood symptoms, which are prevalent in PD^8^, may also affect daily functioning, but it remains unclear whether these effects are independent of those of cognitive impairment, and whether the severity of mood symptoms might confound or mechanistically influence the relationship between cognitive performance and cognition-dependent IADLs.

There are generally two approaches to assessing cognitive function in patients with neurological disorders: 1) performance-based measures, such as standardized neuropsychological tests, and 2) patient-reported outcome measures, such as functional assessments of IADLs like the PDAQ. The PDAQ was developed for PD to isolate cognition-related daily functions from motor-reliant functions^7^.Patient-reported outcome measures serve as important complements, and potentially as more ecologically-valid alternatives to performance-based measures because they capture how cognitive impairments manifest in real-world settings rather than in controlled laboratory environments. However, by capturing everyday function, it is likely that these patient-reported outcome measures also reflect deficits in other domains such as mood^9^. Given the interest in potentially using patient-reported outcome measures in interventional trials targeting cognition, it is crucial to determine the degree to which measures reflect cognition alone or a combination of effects from other symptoms that may not be targeted by the intervention. Indeed, regulatory bodies such as the FDA emphasize construct validity as a fundamental criterion in selecting patient-reported measures^10–12^.

Mood and behavioural symptoms are common in PD patients^13^. Depression affects up to 60% of PD patients^14^, apathy is present in over a third of patients^15^, and anhedonia and impulse control disorders affect nearly half of patients^16,17^. Furthermore, it is well established from studying PD and other populations that mood symptoms impact the ability to perform IADLs^18–21^. In the case of PD, the occurrence and severity of both mood and cognitive symptoms are linked to disease progression^22,23^, raising the possibility that mood symptoms could confound the observed relationship between cognitive impairment and daily function. In older adults, it has also been shown that cognitive impairments may indirectly influence everyday functioning via their effect on mood symptoms^24^. The extent to which mood symptoms contribute to cognitive-dependent IADL impairment in PD, either independently of cognitive function or by confounding or mediating the relationship between cognitive impairment and functional decline, remains unclear.

To address these gaps, we examined a sample of 104 PD patients who had completed the PDAQ to assess cognition-dependent daily function; mood questionnaires to assess apathy, depression, impulsivity, anhedonia; and standard neuropsychological tests of attention/working memory and executive function. First, we aimed to determine whether mood symptoms are significantly associated with cognition-dependent functional ability in PD. Second, we aimed to determine whether mood and cognition independently contribute to cognition-dependent functional ability and to compare the magnitude of their contributions. Third, we aimed to explore whether mood symptoms confound or mediate the relationship between cognitive performance and functional ability. Finally, we replicated the analyses in an online cohort of 144 PD patients who completed an identical set of questionnaires and a similar set of neuropsychological tests.

## Methods and Materials

### Discovery sample

Data for 104 patients with PD were included from a larger in-person cohort of participants from the Quebec Parkinson’s Network (QPN) study, which relied on patients being referred by their neurologist on the basis of a diagnosis of PD^25,26^. To ensure a representative sample there were no exclusion criteria other than having a major co-morbid neurological disorder. Participants were included in the present analysis if they had completed the relevant tasks and questionnaires (see below). Patients underwent the study assessments in their usual medicated state. All participants provided informed consent and received compensation of $25 (CAD). Study assessments were completed by participants in their preferred language (either English or French). The study was approved by the McGill University Health Centre Research Ethics Board.

As not all participants completed all tests, analyses were carried out on varying, but mostly overlapping, subsamples to maximize sample size in each analysis (Supplementary Figure 1).

### Replication sample

Data for 144 patients with PD were selected from a larger online cohort of participants recruited through the Canadian Open Parkinson Network registry^27^ if they had completed the relevant tasks and questionnaires (see below). Recruitment criteria for the parent study included age between 50 and 90 and absence of a diagnosis of dementia based on self-report. Patients were asked to complete the online questionnaires and tasks in their usual medicated state. Participants completed the study in either English or French, according to their preference. All participants were entered into a draw for one of ten $100 (CAD) Amazon e-gift cards. The study was conducted entirely online and received approval from the McGill University Health Centre Research Ethics Board. All participants provided informed consent.

### Performance-Based Measures of Cognition

Participants in the discovery sample completed a standard in-person neuropsychological battery. Executive function was assessed with the Delis-Kaplan Executive Function Color-Word Interference Test (D-KEFS CWIT; response time on the Inhibition condition), Trail Making Test (TMT; the difference in completion time between Part B and Part A in seconds), and the Brixton Spatial Anticipation Test (BSAT; number of errors). Attention/working memory was assessed using Digit Span Test (forward and backward, reverse-coded) and the TMT Part-A (seconds).

Participants in the replication sample completed a set of overlapping but not identical computerized tests, the administration of which has been previously described^28^. Executive function was assessed with TMT (Part B and Part A in seconds), Color-Word Stroop (response time difference on incongruent minus congruent trials in seconds and number of errors). Testing for attention/working memory was identical to the discovery sample.

### Patient-Reported Measures

Participants in both samples completed self-reported questionnaires, including the PDAQ–15, a 15-item questionnaire assessing cognitive daily functioning in PD, with scores ranging from 0-60^29^. The mood questionnaires included the Apathy Evaluation Scale (AES) with scores ranging from 18 to 72^30^; the Geriatric Depression Scale - Short Form (GDS) with scores ranging from 0 to 15^31^; the Questionnaire for Impulsive-Compulsive Disorders in Parkinson’s disease (QUIP) with scores ranging from 0 to 112^32^; the Barratt Impulsiveness Scale-11 (BIS-11) with scores ranging from 30 to 120^33^; and the Snaith Hamilton Pleasure Scale (SHAPS), a questionnaire evaluating anhedonia with scores ranging from 0 to 14^34^. PDAQ was reverse-scored so that, across all measures, higher scores indicated greater impairment. Scores distributions are shown in **Supplementary Figure S1**.

### Statistical Analysis

All analyses were completed in R version 2024.12.0. Significance was determined at p <0.05.

### Independent variables: Cognitive composite scores, Mood questionnaire scores, and Covariates

Performance on each cognitive test was normalized within its respective sample (i.e., discovery or replication). Domain composite scores for Executive Function and Attention/Working Memory were derived by averaging z-scores of tests within each cognitive domain. Mood questionnaires scores were also standardized to allow for comparison of effect sizes between mood and cognition.

Neither age nor education were significantly associated with PDAQ score (β = 0.13, p = 0.239; β = - 0.33, p = 0.303) so they were not retained in subsequent models. Sex was also not associated with PDAQ score (β = -0.32, p = 0.727), but due to the difference in sex distribution within groups, we nonetheless included it as a covariate in all models and additionally examined interactions with sex. Sex was effect-coded, with males as the reference group.

### Regression Analyses

First, we separately modeled the relationship between each mood score and each cognitive composite score with PDAQ score. Then, to test for independent effects of mood and cognition on PDAQ scores we included both in the model. To identify potential confounding effects of mood on the relationship between cognition and PDAQ, we relied on the 10% change-in-estimate criterion^35^.

### Mediation Analyses

We conducted an exploratory mediation analysis using the *mediate* package in R to assess whether mood mediates the effect of cognition on PDAQ scores. The model included cognitive composite score as the independent variable, mood score as the mediating variable, and PDAQ score as the dependent variable. First, the mood score was regressed on the cognitive composite score (e.g., Impulsivity Score ∼ Executive Function). This step assessed whether cognitive performance significantly predicts mood symptom, establishing an initial “a” path relationship necessary for mediation. If the relationship between cognition and mood was significant, the full mediation model was bootstrapped using the percentile method^36^ to evaluate whether the mediator carried a significant portion of the effect of mood score on PDAQ score. A partial mediation effect is present when the indirect effect (Path a × b) is significant, and the direct effect of cognition on PDAQ score (Path c′) is reduced but still significant after including mood as a mediator. A full mediation effect is detected when the indirect effect is significant and the direct effect becomes non-significant, indicating that mood fully accounts for the relationship between cognition and PDAQ score.

## Results

### Sample Characteristics

Demographic and clinical characteristics of the discovery sample are presented in Table 1 and a comparison of the two sample is shown in Supplementary Table 1. The two samples had similar sex distributions and education. Mood questionnaire scores were also similar across samples.

**Table 1.**
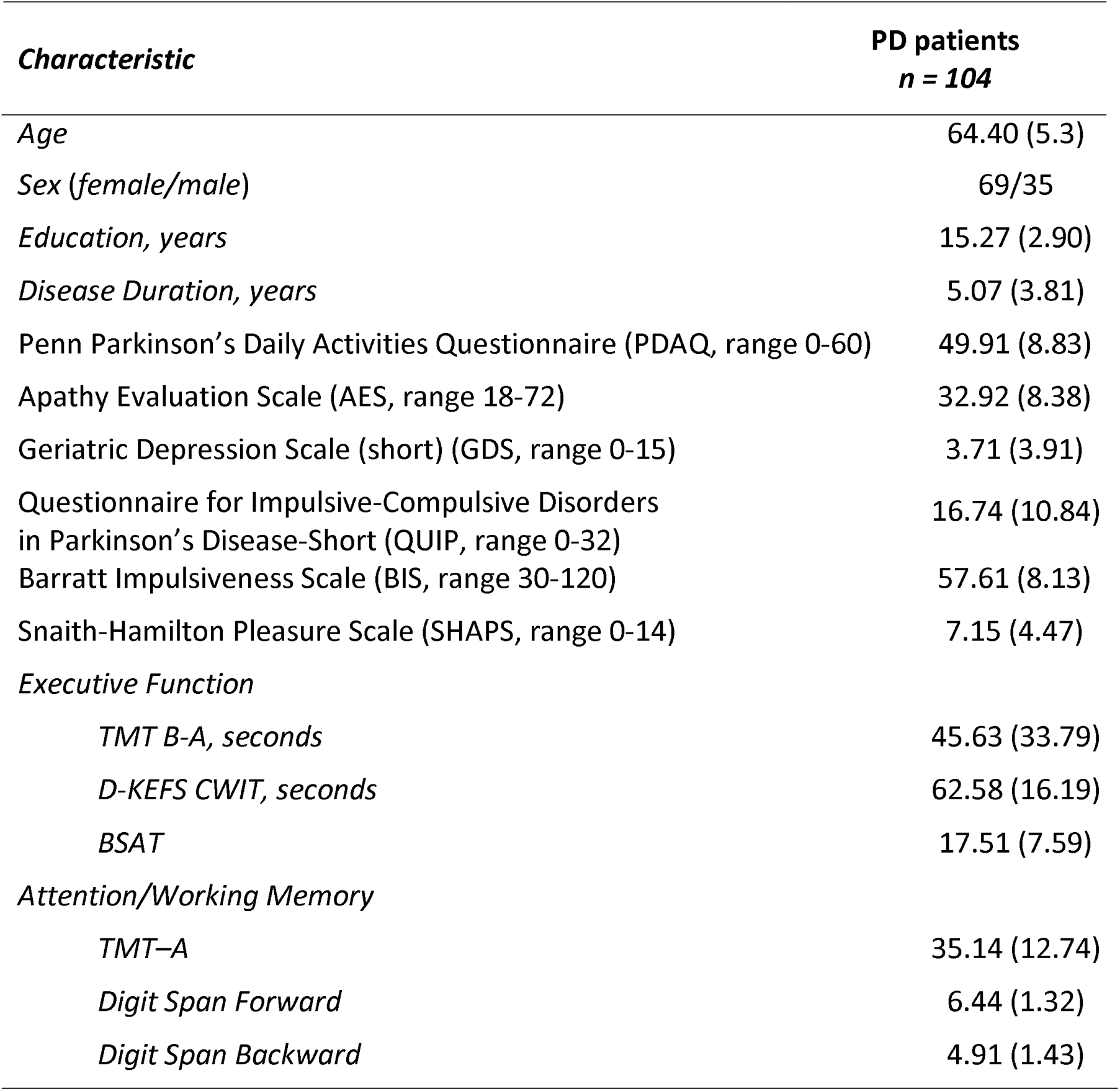
Participant characteristics. Values represent Mean (SD). Abbreviations: Difference in Trail Making Test Part B and Part A (TMT B-A), Trail Making Task Part A (TMT-A), Delis-Kaplan Executive Function (D-KEFS) Color-Word Interference Test (CWIT), Brixton Spatial Anticipation Test (BSAT).

| *Characteristic* | **PD patients**<br>***n* = 104** |
| --- | --- |

|  |  |
| --- | --- |
| *Age* | 64.40 (5.3) |
| *Sex (female/male)* | 69/35 |
| *Education, years* | 15.27 (2.90) |
| *Disease Duration, years* | 5.07 (3.81) |
| Penn Parkinson's Daily Activities Questionnaire (PDAQ, range 0-60) | 49.91 (8.83) |
| Apathy Evaluation Scale (AES, range 18-72) | 32.92 (8.38) |
| Geriatric Depression Scale (short) (GDS, range 0-15) | 3.71 (3.91) |
| Questionnaire for Impulsive-Compulsive Disorders<br>in Parkinson's Disease-Short (QUIP, range 0-32) | 16.74 (10.84) |
| Barratt Impulsiveness Scale (BIS, range 30-120) | 57.61 (8.13) |
| Snaith-Hamilton Pleasure Scale (SHAPS, range 0-14) | 7.15 (4.47) |
| *Executive Function* |  |
| *TMT B-A, seconds* | 45.63 (33.79) |
| *D-KEFS CWIT, seconds* | 62.58 (16.19) |
| *BSAT* | 17.51 (7.59) |
| *Attention/Working Memory* |  |
| *TMT-A* | 35.14 (12.74) |
| *Digit Span Forward* | 6.44 (1.32) |
| *Digit Span Backward* | 4.91 (1.43) |

The replication sample showed moderately higher PDAQ scores (greater impairment, score reversed for consistency). Among cognitive measures, only Digit span performance can be directly compared across samples, as the paper versus computerized format is not expected to meaningfully affect measurement characteristics, and performance was comparable across samples.

### Mood scores and cognitive performance are individually associated with PDAQ score

First, as expected, in the discovery sample, worse executive function (β = 8.79, p < 0.001) and attention/working memory (β = 5.23, p < 0.001) scores were significantly associated with worse PDAQ scores^7,29^. Second, examining the associations between mood and PDAQ, we found that in all cases except for anhedonia (SHAPS: β = 1.67, p = 0.058), worse mood was significantly associated with worse PDAQ score (apathy, AES: β = 3.91, p < 0.001; depression, GDS: β = 4.22, p < 0.001; impulsivity, QUIP: β = 3.91, p < 0.001 and BIS: β = 4.18, p < 0.001) except for anhedonia score. These results are summarized in Figure 1A.

**Figure 1.**
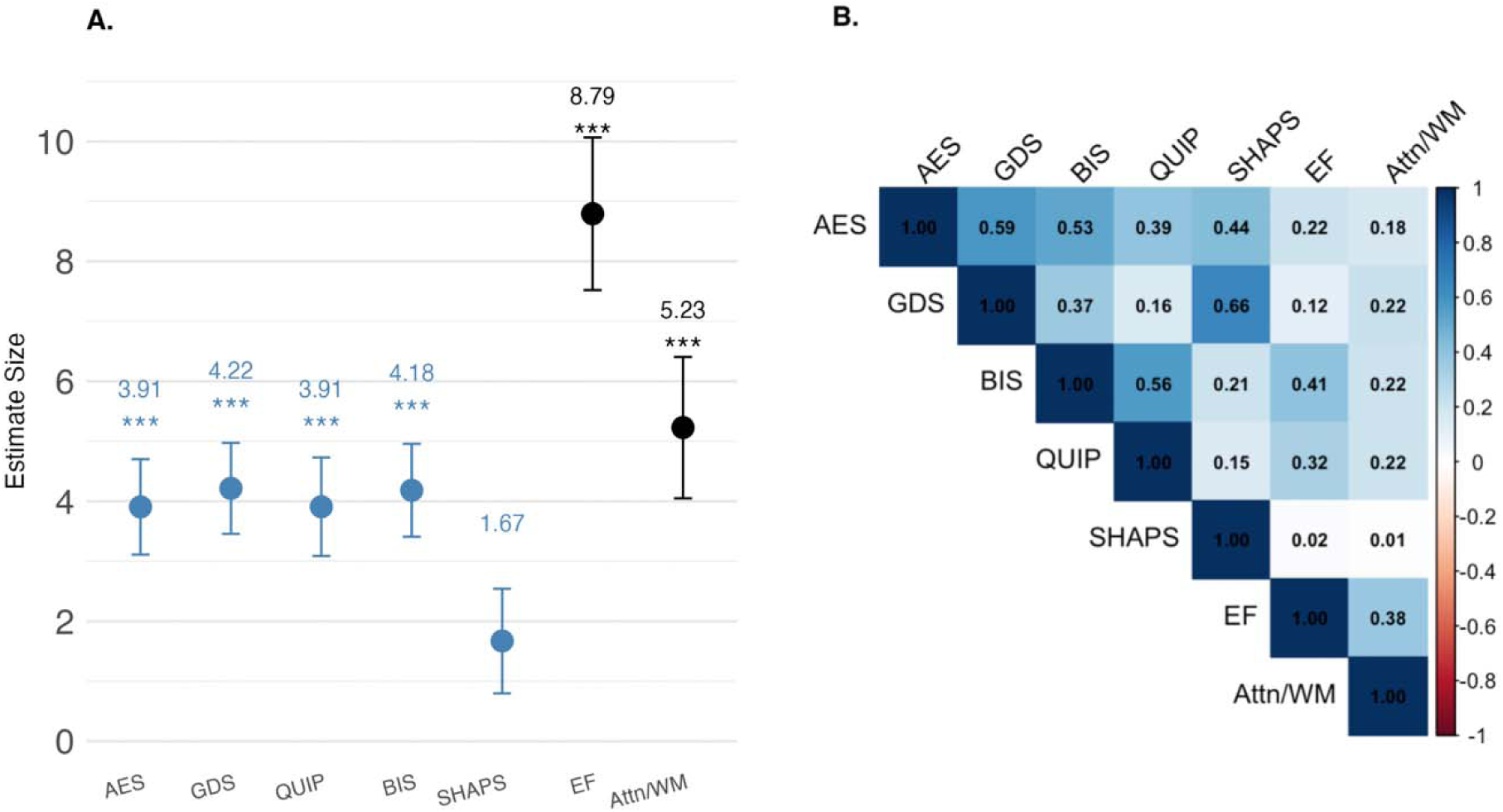
**(A) Standardized beta estimates of the association of mood and cognitive scores with PDAQ.** Regression coefficients (beta estimates) and 95% confidence intervals are shown for each predictor and were each derived from separate linear regression models that controlled for sex. Each predictor was z-scored prior to analysis, allowing effect sizes to be interpreted as the change in PDAQ score associated with a 1 SD change in each predictor. Predictors are color-coded by domain: mood-related (blue) and cognitive (black). **(B) Correlation matrix of mood and cognition predictors.** Pairwise Pearson coefficients are displayed for all mood and cognition predictors included in the regression analyses. Cognitive variables (EF, Attn/WM) show weak to no correlation with mood-related variables (AES, GDS, BIS, QUIP, SHAPS), indicating low collinearity across mood and cognition. Abbreviations: Executive Function (EF), Attention/Working Memory (Attn/WM).

In the replication sample, attention/working memory scores were also significantly associated with PDAQ scores (β = 2.06, p = 0.035), as were all mood scores (AES: β = 5.14, p < 0.001; BIS: β = 5.21, p < 0.001; QUIP: β = 3.84, p < 0.001; GDS: β = 4.13, p < 0.001; SHAPS: β = 2.73, p < 0.001). In contrast, executive function scores were not significantly associated with PDAQ scores (β = –0.87, p = 0.442). Because this was unexpected, we examined the associations between each of the component executive function tests and PDAQ score. TMT B time showed an unexpected association where longer time was associated with better PDAQ scores. Notably, the other subcomponents of the executive function composite score showed the expected associations (see Supplementary Figure 8).

To assess sex effects, we tested sex-by-predictor interactions in both samples (Supplementary Figure 3) and found no significant interactions. Therefore, interactions were not included in subsequent models, but sex was retained as a covariate.

### Examining the independent contributions of mood and cognition to PDAQ score

First, we examined pairwise correlations to ensure that mood and cognitive scores were not strongly correlated. In the discovery sample, mood-mood correlations were weak to moderate (r = 0.15–0.66) and mood-cognition correlations were weak (r = 0.01–0.41; Figure 1B). Similar patterns were observed in the replication sample (Supplementary Figure 5).

We then examined whether including both mood scores and cognitive performance in the same model improved explained variance compared to models with cognitive performance alone. In the discovery sample, adding mood scores to the executive function model increased the adjusted R² by 0.02 to 0.13 compared to the base executive function model (R_base_² = 0.34; Figure 2A), representing a 6 to 39% increase in explained PDAQ variance. Adding mood scores to the attention/working memory model increased the adjusted R² by 0.03 to 0.21 compared to the base model (adjusted R_base_² = 0.16; Figure 2B), representing 62 to 177% in explained variance.

**Figure 2.**
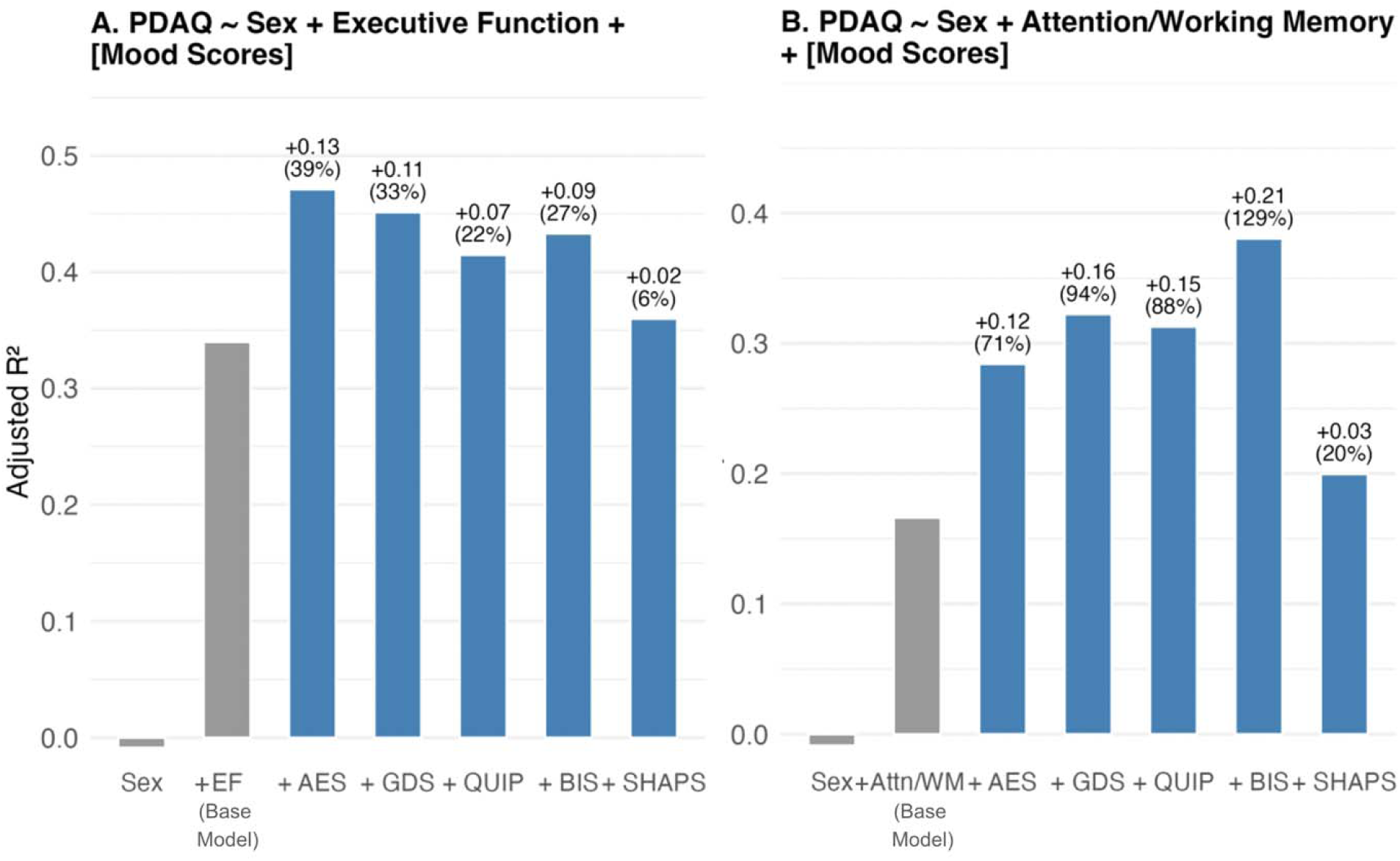
Comparison of variance explained in cognition-related daily functioning (PDAQ) by adding mood scores to base cognitive models. **(A)** Models including sex alone, then sex and executive function (EF), and then with the addition of each individual mood variable. **(B)** Models including sex alone, then sex and attention/working memory (Attn/WM), and then with the addition of each individual mood variable. Each bar represents the adjusted R² of the linear regression model. Text labels above each bar represent the absolute and relative increase in explained variance in models that add each of the mood measures compared to the base model that includes the cognitive measure and sex only.

Importantly, in all cases, when both were included in the models, cognitive and mood scores remained significant predictors of PDAQ score (Figure 3). Results were similar in the replication sample (Supplementary Fig 7). These findings suggest that overall, mood scores contribute significant additional, independent variance to PDAQ scores, and, in the case of the relationship between attention/working memory and PDAQ, mood contributes a similar magnitude of explained variance.

**Figure 3.**
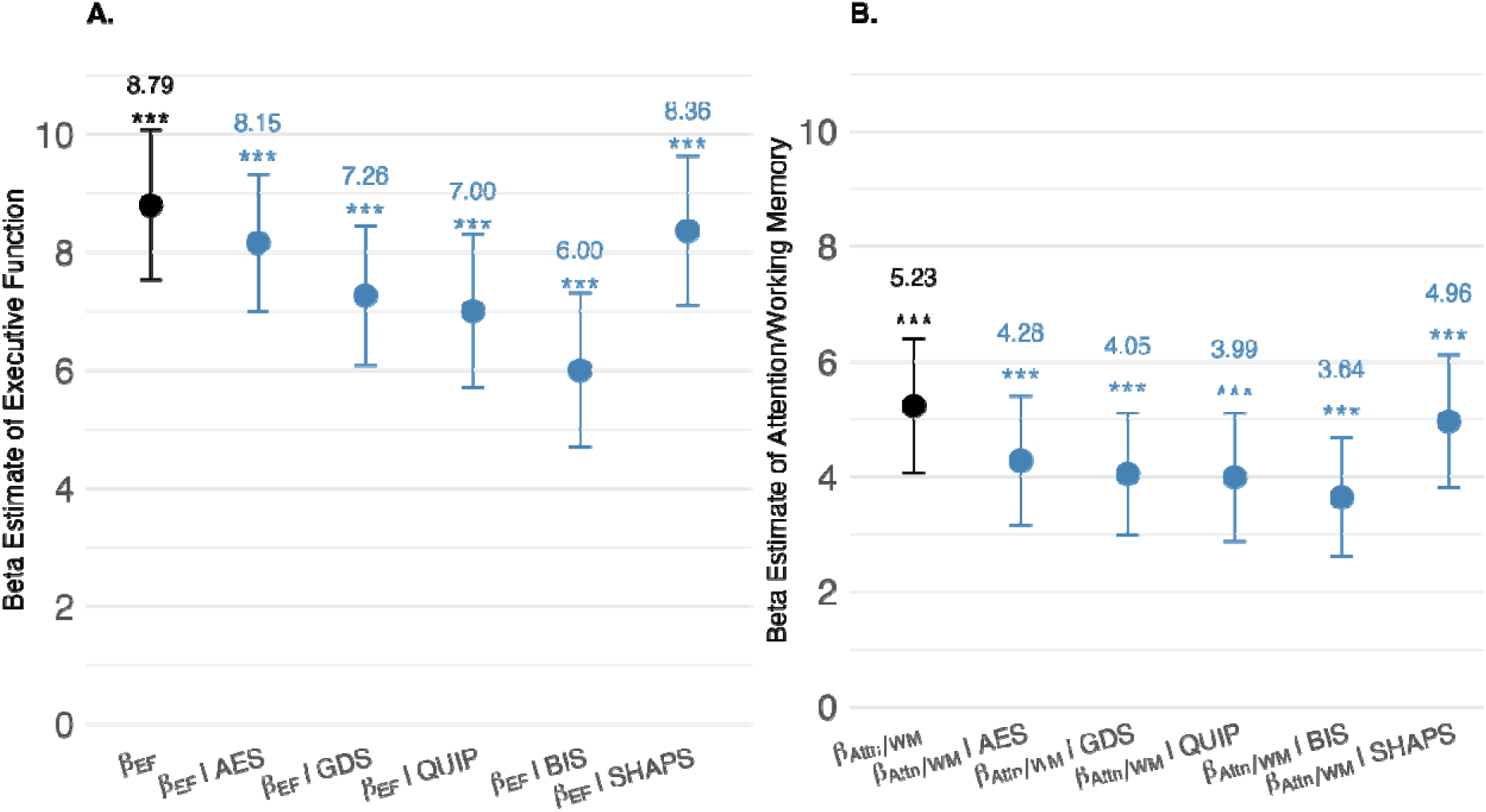
Assessment of confounding of the relationship between cognition and cognitive IADLs (PDAQ) by different mood symptoms. **(A)** Beta estimates for the effect of executive function (EF) on PDAQ while controlling for different mood predictors (light blue) compared the beta estimate of for the effect of EF when mood score is not included in the model (dark blue). **(B)** Beta estimates for the effect of attention/working memory (Attn/WM) on PDAQ while controlling for mood predictors (light blue) compared the beta estimate of for the effect of Attn/WM when mood score is not included in the mode (dark blue). In both models, all variables were z-scored to allow standardized effect size comparisons. Error bars represent 95% confidence intervals.

To determine whether mood symptoms are confounding the relationship between cognitive performance and PDAQ score, we compared the estimate sizes for the relationship between cognition and PDAQ in the models that did and did not additionally include the mood scores. In the case of executive function, the addition of depression (GDS: β_EF_ = 8.36, p <0.001, 17.4 % reduction in β), and of impulsivity scores (QUIP: β_EF_ = 7.00, p <0.001, 20.4 % reduction in β; BIS: β_EF_ = 6.00, p <0.001, 31.7% reduction in β) led to a reduction in the effect size of executive function on PDAQ score by more than 10%, meeting the criterion for confounding (Figure 3A), whereas the addition of apathy (AES: β_EF_ = 8.15, p <0.001, 7.3% reduction in β) and anhedonia (SHAPS: β_EF_ = 7.01, p <0.001, 4.9% reduction in β) did not. In the case of Attention/working memory, the addition of apathy (AES: β_Attn/WM_ = 4.28, p <0.001, 18.2% reduction in β), depression (GDS: β_Attn/WM_= 4.05, p <0.001, 22.6% reduction in β), and impulsivity (QUIP: β_Attn/WM_= 3.99, p <0.001, 23.7% reduction in β ; BIS: β_Attn/WM_ = 3.64, p <0.001, 30.4% reduction in β) led to a reduction in the effect size of attention/working memory on PDAQ score by more than 10% (Figure 3B), meeting the criterion for confounding, whereas adjustment for anhedonia (SHAPS: β_Attn/WM_ = 8.36, p <0.001, 5.2% reduction in β) did not.

In the replication sample, because executive function was not significantly associated with PDAQ score, no further analyses were conducted for this domain. Analyses with the attention/working memory domain showed similar results to those in the discovery sample, except that depression and anhedonia did not meet threshold for confounding (Supplementary Figure 4).

### Mood significantly mediates the relationship between cognition and PDAQ score

Finally, we examined whether mood symptoms mediated the relationship between cognition and PDAQ score. In the discovery sample, significant associations were observed between executive function and the impulsivity mediators QUIP and BIS, warranting further mediation analyses. These models demonstrated a partial mediation by impulsivity symptoms as evidenced by both significant direct (c′) and indirect (a*b) effects (Figure 4). No mediation analyses were conducted for GDS, SHAPS, nor AES, as the association between executive function and these mediators was not significant.

**Figure 4.**
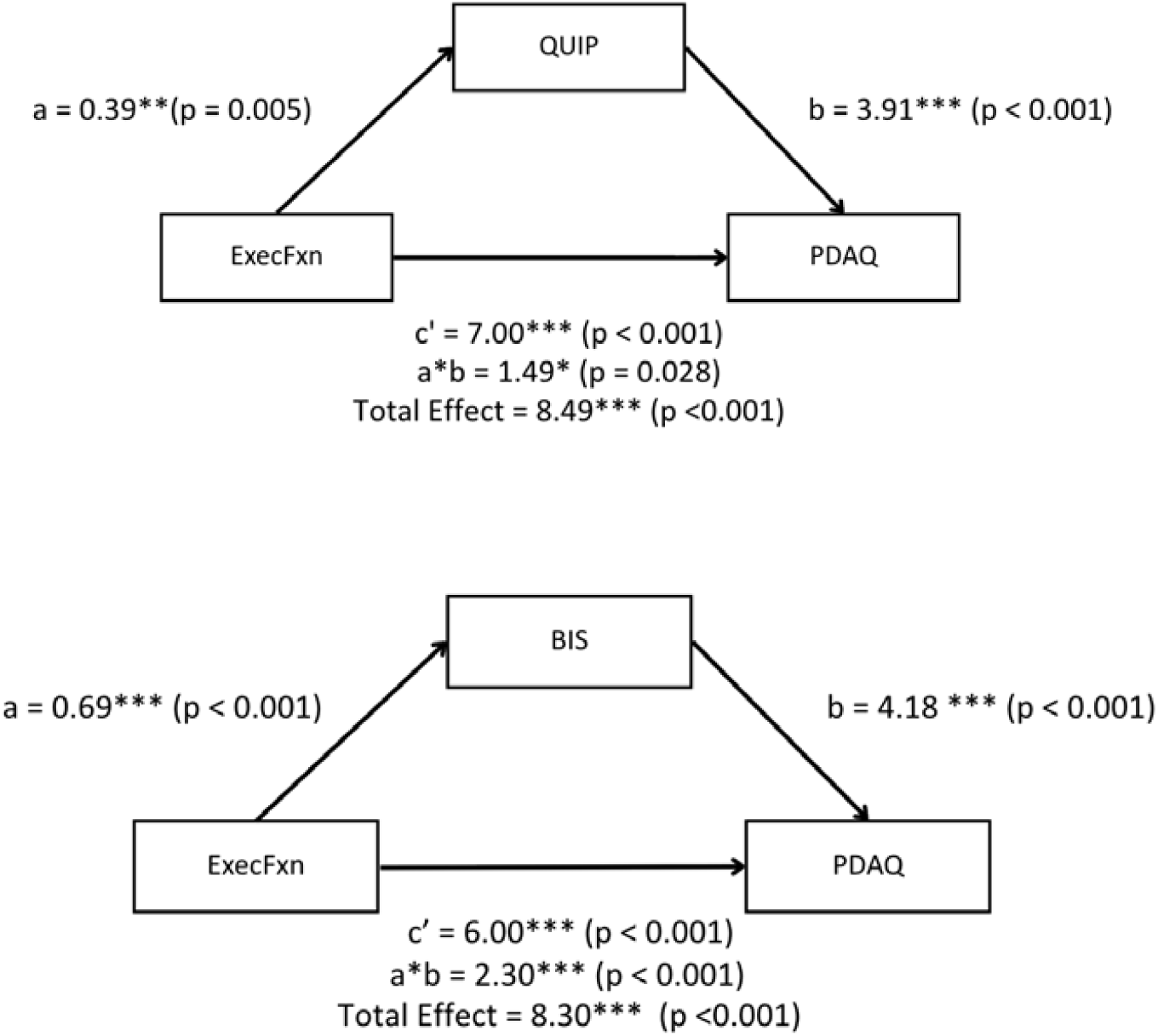
Mediation models testing whether mood symptoms mediate the effect of executive function (ExecFxn) on cognitive IADLs (PDAQ). Each panel shows standardized path coefficients (β) and p-values for the “a” path (ExecFxn → Mood), “b” path (Mood → PDAQ), and “c’” path (ExecFxn → PDAQ, controlling for the mediator). The total effect represents the overall association between ExecFxn and PDAQ before accounting for the mediator. All models include sex as a covariate. Significant indirect effects were observed for BIS (impulsivity), and QUIP (compulsivity), indicating partial mediation of the relationship between executive function and functional outcome.

Additionally, no significant relationships between attention/working memory and mood symptoms were found, neither in the discovery nor the replication sample, precluding further analyses for this domain.

## Discussion

The goal of this study was to measure the degree to which mood symptoms contribute to the impairment in cognitive IADLs, and whether these contributions are independent of, and comparable in magnitude to those of cognitive deficits. We found that each of the mood symptoms assessed was independently associated with cognitive IADL function, and, that in some cases, the degree of explained variance in IADL function was similar for mood and cognitive performance. Among mood symptoms, only impulsivity partially mediated the relationship between worse cognitive performance and greater impairment in cognitive IADLs. This suggests that, except for impulsivity, largely independent mechanisms link mood and cognitive symptoms to functional impairment. Most findings were replicated in an independent sample. Our findings suggest that mood symptoms are a significant independent contributor to cognition-dependent functional impairment in PD and that the presence of mood symptoms should be taken into careful consideration when designing interventional studies that use a measurement of cognitive IADLs as the outcome measure.

We found that mood symptoms (apathy, depression, and impulsivity) were significantly and independently associated with cognitive IADL impairment in individuals with PD, with a relative increase in explained variance raging from 20-129%. This finding is consistent with previous research in aging populations and in Alzheimer’s disease, where mood symptoms such as depression have been shown to predict IADL deficits^37^. In PD, where mood symptoms are prevalent and considered a direct consequence of neurodegeneration^14,38–40^, the role of mood in IADLs has been much less explored. In the original validation study of the PDAQ, a tool specifically designed to assess cognitive IADLs in PD and therefore proposed as a potential PRO for tracking the functional benefits of cognitive-enhancing interventions, the authors addressed concerns about mood-related confounds by demonstrating that depression was not associated with PDAQ scores. In contrast, in the present study, we found, in two independent samples, that depression, as well as apathy and impulsivity, were independently associated with PDAQ scores after accounting for cognitive performance. One possible explanation for these discordant results is that our patient sample was less cognitively impaired than the patients included in the development and validation of the PDAQ, which included patients with dementia^7,29^. Depression levels, however, were comparable to ours (mean GDS score of 3.6 compared to 3.7 and 4 in our discovery and replication samples, respectively). This suggests that the PDAQ might be better suited to measure more severe cognitive impairment, a stage when mood symptoms may play a relatively more minor role in cognition-dependent functional impairments. However, the PDAQ has also been shown to be more sensitive than neuropsychological testing for capturing subjective cognitive impairment – the earliest stage of cognitive dysfunction^41^, which is arguably the stage at which sensitive and specific outcome measures are most needed. Our results suggest that use of the PDAQ in the earlier stages of the disease, when patients are minimally impaired, also requires measurement of mood symptoms to account for their independent effects.

Because neuropsychiatric symptoms can covary with cognitive impairments in PD^8,22^,we evaluated for both confounding and mediation. Confounding analyses evaluate whether the observed relationship between cognitive performance and daily function is biased by a shared relationship to neuropsychiatric symptoms. Mediation analyses, in contrast, evaluate whether neuropsychiatric symptoms could represent a mechanistic pathway through which cognitive dysfunction leads to functional impairments. Evaluating both allows for distinguishing explanations that rely on shared variance from those that rely on the existence of a mechanistic pathway. We found evidence for both. First, our results suggest that higher depression and higher impulsivity partially confound the relationship between cognitive impairment and cognitive IADLs. This implies that cognitive IADL impairments previously attributed to cognitive dysfunction are partly accounted for by overlapping effects of depression and impulsivity symptoms on IADLs. We also found that higher impulsivity (but not depression nor other symptoms) partially mediated the relationship between executive dysfunction and IADL impairments. This suggests a possible indirect behavioral pathway whereby executive deficits contribute to impaired self-regulation, which could then undermine everyday task execution. One possible explanation for the presence of both confounding and mediation is shared neural circuitry within dopaminergic frontostriatal networks contributing to both cognitive dysfunction, and executive control in particular, and neuropsychiatric symptoms. Degeneration affecting partially overlapping cognitive (dorsolateral prefrontal–dorsal caudate) and limbic (orbitofrontal/anterior cingulate–ventral striatum) loops could amplify the coupling between cognitive and behavioural dysfunction in PD^42–44^.

Our findings show that mood symptoms and cognitive deficits independently contribute to cognitive IADL impairment in PD. The mechanisms underlying these relationships are likely distinct. Whereas executive dysfunction and attentional impairments are thought to directly compromise everyday task performance (e.g., impaired planning affects medication management)^2–4^, neuropsychiatric symptoms likely affect IADL performance through motivational pathways. Apathy, for example, has been shown in Alzheimer’s disease and Mild Cognitive Impairment to disrupt IADLs by reducing the ability to initiate and sustain activities^45^, which could also apply to PD. Depression and anhedonia may impair functioning by reducing engagement in activities, potentially causing a self-reinforcing cascade of inactivity and further functional decline^46,47^. Impulsivity may impair IADLs through reduced inhibitory control and poor monitoring of negative behaviors^48^. The possibility of distinct functional mechanisms underlying the relationship between neuropsychiatric symptoms and daily functioning further underscores the importance of considering these symptoms when measuring and developing interventions for functional impairments.

A few limitations warrant consideration. First, the absence of anxiety and psychosis measures leaves gaps in our understanding of how other common neuropsychiatric symptoms in PD influence functional outcomes^8^. Second, the shared self-report methodology for PDAQ and mood questionnaires raises the possibility of inflated associations due to common method variance, rather than true mechanistic links^49^. Future work could consider including knowledgeable informant-reported data (e.g., caregiver ratings)^50^. Third, most but not all results were replicated in the replication sample, in particular owing to an unexpected direction for the relationship between TMT B performance and IADL impairment. Importantly, however, the significant and independent relationships between neuropsychiatric symptoms and cognitive IADL performance were replicated across both samples demonstrating the robustness of the findings.

In conclusion, we found that neuropsychiatric symptoms are strong predictors of cognitive IADL impairments as measured by the PDAQ, and that their effect is independent of that of cognitive deficits. The PDAQ is the first disease-specific measure of cognitive-dependent instrumental activities of daily living in PD and shows potential as a patient-reported outcome measure for cognitive intervention trials. Our results suggest that if such a measure is to be used, mood symptoms should be broadly assessed and explicitly controlled for to determine whether intervention-related functional improvements are driven by direct cognitive enhancement, mood alleviation or both.

## Data Availability

All data produced in the present study are available upon reasonable request to the authors

## Acknowledgments

Yilin Zhang, Nasri Balit, Sarah Bogard, Roozbeh Sattari, Courtney Jackson, Lucy Zou for their help with data collection, QPN and COPN for their help with recruitment, and all participants for their time and commitment to research.

This research was supported by a grant from the Canada First Research Excellence Fund-Healthy Brains for Healthy Lives and from the Canadian Institute for Health Research (MS), a salary award from the Fonds de Recherche du Quebec – Santé (MS), a graduate student award from Parkinson Canada (SS) and an undergraduate student award from the National Sciences and Engineering Research Council of Canada (JYH).

## Author contributions

Jun Yu Hu: Conceptualization, Formal analysis, Investigation, Methodology, Writing – original draft. Sophie Sun: Conceptualization, Investigation, Methodology, Writing – review & editing. Madeleine Sharp: Conceptualization, Project administration, Resources, Supervision, Writing – review & editing.

## References

1. Goldman, J. G. et al. Cognitive impairment in Parkinson’s disease: a report from a multidisciplinary symposium on unmet needs and future directions to maintain cognitive health. Npj Park. Dis. 4, 1–11 (2018).

2. Becker, S. et al. Cognitive-Driven Activities of Daily Living Impairment as a Predictor for Dementia in Parkinson Disease: A Longitudinal Cohort Study. Neurology 99, (2022).

3. Foster, E. R. & Doty, T. Cognitive Correlates of Instrumental Activities of Daily Living Performance in Parkinson Disease Without Dementia. Arch. Rehabil. Res. Clin. Transl. 3, 100138 (2021).

4. Pirogovsky, E. et al. Instrumental activities of daily living are impaired in Parkinson’s disease patients with mild cognitive impairment. Neuropsychology 28, 229–237 (2014).

5. Cahn, D. A. et al. Differential Contributions of Cognitive and Motor Component Processes to Physical and Instrumental Activities of Daily Living in Parkinson’s Disease. Arch. Clin. Neuropsychol. 13, 575–583 (1998).

6. Katz, S. Assessing Self-maintenance: Activities of Daily Living, Mobility, and Instrumental Activities of Daily Living. J. Am. Geriatr. Soc. 31, 721–727 (1983).

7. Brennan, L. et al. Development and initial testing of the Penn Parkinson’s Daily Activities Questionnaire. Mov. Disord. 31, 126–134 (2016).

8. Weintraub, D. et al. The neuropsychiatry of Parkinson’s disease: advances and challenges. Lancet Neurol. 21, 89–102 (2022).

9. Numbers, K. et al. Instrumental Activities of Daily Living by Subjective and Objective Measures: The Impact of Depression and Personality. Front. Aging Neurosci. 14, 829544 (2022).

10. Kaneyasu, T. et al. How to select and understand guidelines for patient-reported outcomes: a scoping review of existing guidance. BMC Health Serv. Res. 24, 334 (2024).

11. Reeve, B. B. et al. ISOQOL recommends minimum standards for patient-reported outcome measures used in patient-centered outcomes and comparative effectiveness research. Qual. Life Res. 22, 1889–1905 (2013).

12. U.S. Food and Drug Administration. Guidance for Industry: Patient-Reported Outcome Measures: Use in Medical Product Development to Support Labeling Claims. Cent. Drug Eval. Res. Cent. Biol. Eval. Res. Cent. Devices Radiol. Health US Dep. Health Hum. Serv. https://www.fda.gov/regulatory-information/search-fda-guidance-documents/patient-reported-outcome-measures-use-medical-product-development-support-labeling-claims (2009).

13. Bock, M. A. et al. Cognitive and Functional Trajectories in Older Adults With Prediagnostic Parkinson Disease. Neurology 100, e1386–e1394 (2023).

14. Dlay, J. K. et al. Progression of Neuropsychiatric Symptoms over Time in an Incident Parkinson’s Disease Cohort (ICICLE-PD). Brain Sci. 10, 78 (2020).

15. Santangelo, G. et al. Apathy in Parkinson’s disease: diagnosis, neuropsychological correlates, pathophysiology and treatment. Behav. Neurol. 27, 501–513 (2013).

16. Reichmann, H., Brecht, M. H., Köster, J., Kraus, P. H. & Lemke, M. R. Pramipexole in routine clinical practice: a prospective observational trial in Parkinson’s disease. CNS Drugs 17, 965–973 (2003).

17. Corvol, J.-C. et al. Longitudinal analysis of impulse control disorders in Parkinson disease. Neurology 91, e189–e201 (2018).

18. Lee, S. Perceived Health, Psychological Distress, and Subjective Well-Being among Older Adults with Parkinson’s Disease: A Cross-Lagged Analysis. Int. J. Environ. Res. Public. Health 18, 12566 (2021).

19. Randver, R. Repetitive transcranial magnetic stimulation of the dorsolateral prefrontal cortex to alleviate depression and cognitive impairment associated with Parkinson’s disease: A review and clinical implications. J. Neurol. Sci. 393, 88–99 (2018).

20. Numbers, K. et al. Instrumental Activities of Daily Living by Subjective and Objective Measures: The Impact of Depression and Personality. Front. Aging Neurosci. 14, 829544 (2022).

21. Zhao, L., Wang, J., Deng, H., Chen, J. & Ding, D. Depressive Symptoms and ADL/IADL Disabilities Among Older Adults from Low-Income Families in Dalian, Liaoning. Clin. Interv. Aging 17, 733–743 (2022).

22. Riedel, O. et al. Frequency of dementia, depression, and other neuropsychiatric symptoms in 1,449 outpatients with Parkinson’s disease. J. Neurol. 257, 1073–1082 (2010).

23. Weintraub, D. et al. Neuropsychiatric symptoms and cognitive abilities over the initial quinquennium of Parkinson disease. Ann. Clin. Transl. Neurol. 7, 449–461 (2020).

24. Wu, C. The mediating and moderating effects of depressive symptoms on the prospective association between cognitive function and activities of daily living disability in older adults. Arch. Gerontol. Geriatr. 96, 104480 (2021).

25. Gan-Or, Z. et al. The Quebec Parkinson Network: A Researcher-Patient Matching Platform and Multimodal Biorepository. J. Park. Dis. 10, 301–313 (2020).

26. Sun, S. et al. Selective Effects of Substantia Nigra and Locus Coeruleus Degeneration on Cognition in Parkinson’s Disease. Mov. Disord. 40, 844–854 (2025).

27. Cressatti, M. et al. Advancing Parkinson’s Disease Research in Canada: The Canadian Open Parkinson Network (C-OPN) Cohort. J. Park. Dis. 14, 1481–1494 (2024).

28. Balit, N., Sun, S., Zhang, Y. & Sharp, M. Online unsupervised performance-based cognitive testing: A feasible and reliable approach to scalable cognitive phenotyping of Parkinson’s patients. Parkinsonism Relat. Disord. 129, 107183 (2024).

29. Brennan, L. et al. The Penn Parkinson’s Daily Activities Questionnaire-15: Psychometric properties of a brief assessment of cognitive instrumental activities of daily living in Parkinson’s disease. Parkinsonism Relat. Disord. 25, 21–26 (2016).

30. Marin, R. S., Biedrzycki, R. C. & Firinciogullari, S. Reliability and validity of the Apathy Evaluation Scale. Psychiatry Res. 38, 143–162 (1991).

31. Sheikh, J. I. & Yesavage, J. A. Geriatric Depression Scale (GDS): Recent evidence and development of a shorter version. Clin. Gerontol. J. Aging Ment. Health 5, 165–173 (1986).

32. Weintraub, D. et al. Validation of the questionnaire for impulsive-compulsive disorders in Parkinson’s disease. Mov. Disord. Off. J. Mov. Disord. Soc. 24, 1461–1467 (2009).

33. Patton, J. H., Stanford, M. S. & Barratt, E. S. Factor structure of the Barratt impulsiveness scale. J. Clin. Psychol. 51, 768–774 (1995).

34. Snaith, R. P. et al. A scale for the assessment of hedonic tone the Snaith-Hamilton Pleasure Scale. Br. J. Psychiatry J. Ment. Sci. 167, 99–103 (1995).

35. Greenland, S. & Morgenstern, H. Confounding in health research. Annu. Rev. Public Health 22, 189–212 (2001).

36. Pesigan, I. J. A. & Cheung, S. F. SEM-Based Methods to Form Confidence Intervals for Indirect Effect: Still Applicable Given Nonnormality, Under Certain Conditions. Front. Psychol. 11, (2020).

37. Votruba, K. L., Persad, C. & Giordani, B. Patient Mood and Instrumental Activities of Daily Living in Alzheimer Disease: Relationship Between Patient and Caregiver Reports. J. Geriatr. Psychiatry Neurol. 28, 203–209 (2015).

38. Assogna, F., Cravello, L., Caltagirone, C. & Spalletta, G. Anhedonia in Parkinson’s disease: A systematic review of the literature. Mov. Disord. 26, 1825–1834 (2011).

39. Weintraub, D. & Mamikonyan, E. The Neuropsychiatry of Parkinson Disease: A Perfect Storm. Am. J. Geriatr. Psychiatry 27, 998–1018 (2019).

40. González-Usigli, H. A. et al. Neurocognitive Psychiatric and Neuropsychological Alterations in Parkinson’s Disease: A Basic and Clinical Approach. Brain Sci. 13, 508 (2023).

41. Purri, R. et al. Subjective Cognitive Complaint in Parkinson’s Disease Patients With Normal Cognition: Canary in the Coal Mine? Mov. Disord. Off. J. Mov. Disord. Soc. 35, 1618–1625 (2020).

42. Lewis, S. J. G., Dove, A., Robbins, T. W., Barker, R. A. & Owen, A. M. Cognitive Impairments in Early Parkinson’s Disease Are Accompanied by Reductions in Activity in Frontostriatal Neural Circuitry. J. Neurosci. 23, 6351–6356 (2003).

43. Brown, G. et al. Frontostriatal and limbic contributions to cognitive decline in Parkinson’s disease. J. Neuroimaging Off. J. Am. Soc. Neuroimaging 33, 121–133 (2023).

44. Wen, M. -C., Chan, L. L., Tan, L. C. S. & Tan, E. K. Depression, anxiety, and apathy in Parkinson’s disease: insights from neuroimaging studies. Eur. J. Neurol. 23, 1001–1019 (2016).

45. Boyle, P. A. et al. Executive dysfunction and apathy predict functional impairment in Alzheimer disease. Am. J. Geriatr. Psychiatry Off. J. Am. Assoc. Geriatr. Psychiatry 11, 214–221 (2003).

46. Covinsky, K. E., Cenzer, I. S., Yaffe, K., O’Brien, S. & Blazer, D. G. Dysphoria and anhedonia as risk factors for disability or death in older persons: implications for the assessment of geriatric depression. Am. J. Geriatr. Psychiatry Off. J. Am. Assoc. Geriatr. Psychiatry 22, 606–613 (2014).

47. Penninx, B. W., Leveille, S., Ferrucci, L., van Eijk, J. T. & Guralnik, J. M. Exploring the effect of depression on physical disability: longitudinal evidence from the established populations for epidemiologic studies of the elderly. Am. J. Public Health 89, 1346–1352 (1999).

48. Dalley, J. W. & Robbins, T. W. Fractionating impulsivity: neuropsychiatric implications. Nat. Rev. Neurosci. 18, 158–171 (2017).

49. Enkavi, A. Z. et al. Large-scale analysis of test–retest reliabilities of self-regulation measures. Proc. Natl. Acad. Sci. 116, 5472–5477 (2019).

50. Deck, B. L. et al. Cognitive Functional Abilities in Parkinson’s Disease: Agreement Between Patients and Informants. Mov. Disord. Clin. Pract. 6, 440–445 (2019).

