## Supplemental Data for "Shared and distinct effects of cognitive performance and mood on cognition-dependent instrumental activities of daily living in Parkinson’s disease"

### Supplement

| **Characteristic** | **Discovery**  **n = 104** | **Replication**  **n = 144** | **p-value** |
| --- | --- | --- | --- |
| Age | 64.40 (8.05) | 66.36 (7.64) | 0.058 |
| Sex (female/male) | 35/69 | 58/85 | 0.3 |
| Education, years | 15.27 (2.90) | 15.81 (3.44) | 0.2 |
| Disease Duration, years | 5.07 (3.81) | 6.83 (5.19) | 0.005 |
| Penn Parkinson’s Daily Activities Questionnaire | 49.91 (8.83) | 47.24 (8.56) | 0.018 |
| Apathy Evaluation Scale (AES) | 32.92 (8.38) | 31.31 (8.27) | 0.14 |
| Geriatric Depression Scale (short) (GDS) | 3.71 (3.91) | 4.00 (3.49) | 0.5 |
| Questionnaire for Impulsive-Compulsive Disorders in Parkinson’s Disease (short) (QUIP) | 16.74 (10.84) | 15.19 (10.22) | 0.3 |
| Barratt Impulsiveness Scale (BIS) | 57.61 (8.13) | 58.74 (8.71) | 0.3 |
| Snaith-Hamilton Pleasure Scale (SHAPS) | 7.15 (4.47) | 7.22 (4.49) | > 0.9 |
| Executive Function |  |  |  |
| TMT B-A, seconds (discovery) | 45.63 (33.79) |  |  |
| D-KEFS CWIT, seconds (discovery) | 62.58 (16.19) |  |  |
| BSAT | 17.51 (7.59) |  |  |
| TMT B-A, seconds (replication) |  | 34.38 (42.48) |  |
| Color-Word Stroop Incongruent RT |  | 1.04 (0.25) |  |
| Color-Word Stroop Incongruent Error |  | 5.96 (8.27) |  |
| Attention/Working Memory |  |  |  |
| TMT–A (Discovery) | 35.14 (12.74) |  |  |
| TMT—A (Replication) |  | 54.51 (16.08) |  |
| Digit Span Forward | 6.44 (1.32) | 6.10 (1.29) | 0.066 |
| Digit Span Backward | 4.91 (1.43) | 5.03 (1.54) | 0.6 |

**Supplementary Table 1**. Participant characteristics of discovery and replication sample. Values represent Mean (SD). P-value computed by Welch Two Sample t-test; Pearson’s Chi-squared test. Abbreviations: Difference in Trail Making Test Part B and Part A (TMT B-A), Trail Making Task Part A (TMT-A), Delis-Kaplan Executive Function (D-KEFS) Color-Word Interference Test (CWIT), Brixton Spatial Anticipation Test (BSAT).

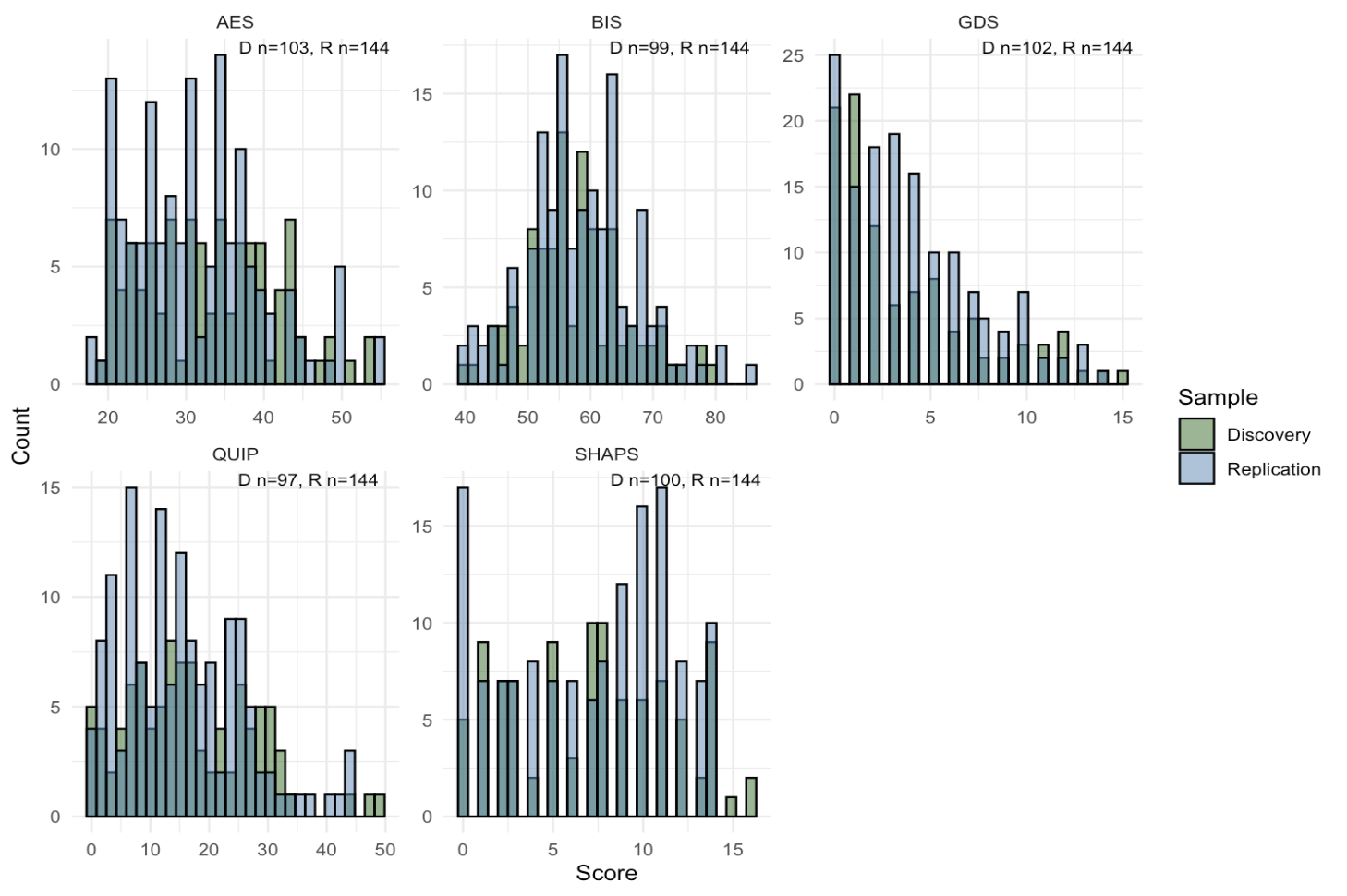

**Supplementary Figure 1**. Distribution of neuropsychiatric questionnaire scores across discovery and replication samples. Histograms show the raw score distributions for the Apathy Evaluation Scale (AES), Barratt Impulsiveness Scale-11 (BIS), Geriatric Depression Scale – Short Form (GDS), Questionnaire for Impulsive-Compulsive Disorders in Parkinson’s Disease (QUIP) – Short, and the Snaith–Hamilton Pleasure Scale (SHAPS). Data are stratified by discovery (green) and replication (blue) cohorts. Sample sizes for each measure are indicated at the top of each panel (D = Discovery, R = Replication). Panels are scaled independently to reflect the distributional properties of the respective measure.

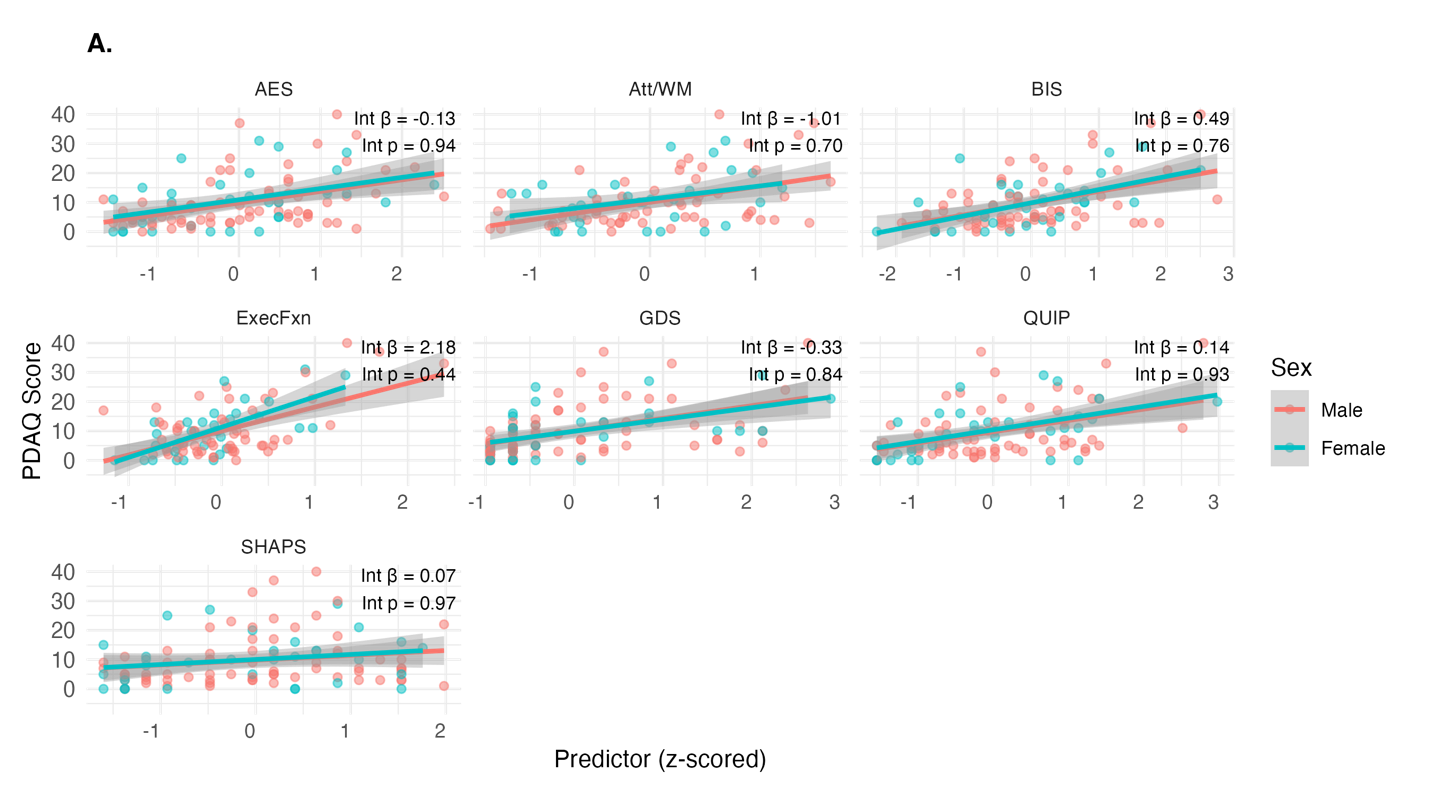

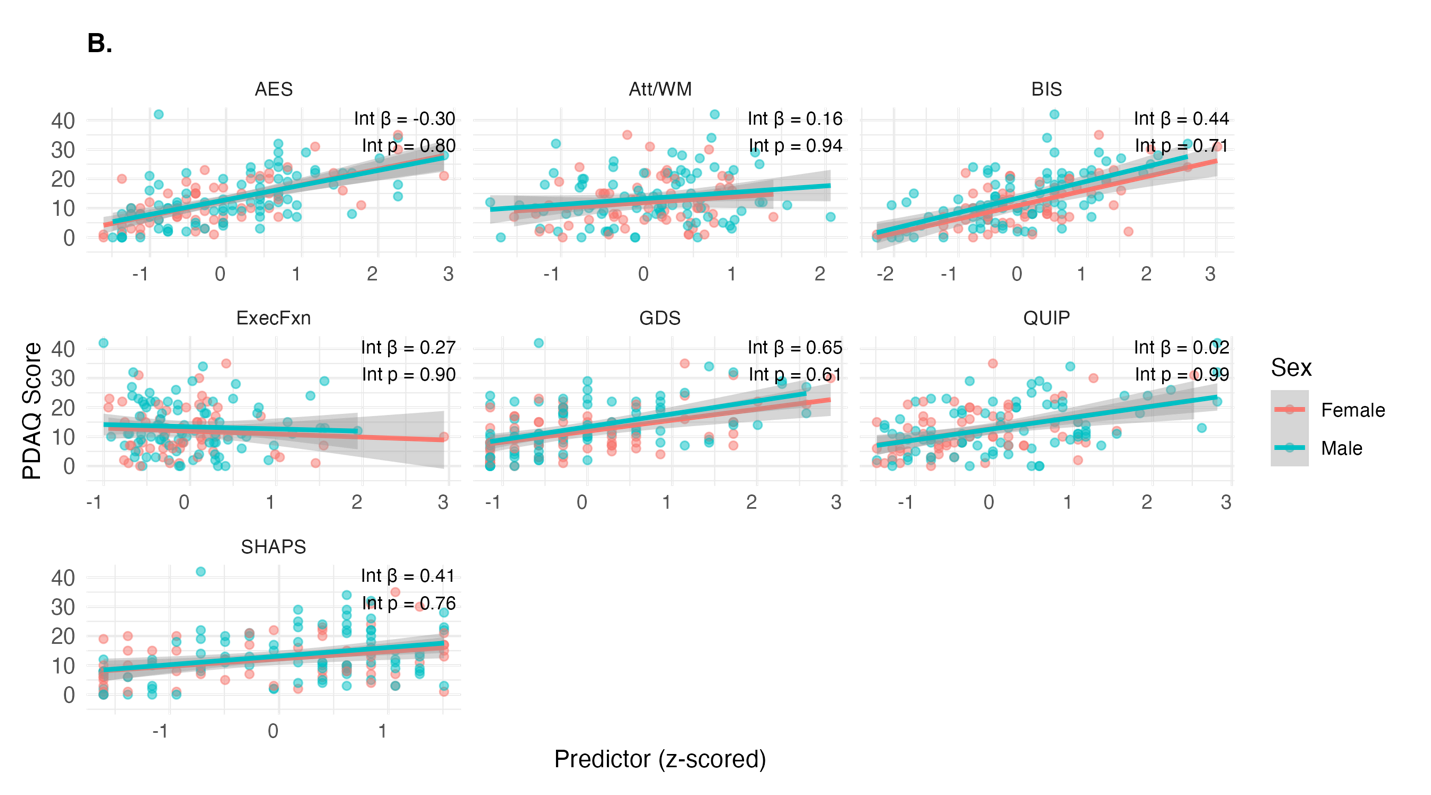

**Supplementary Figure 3. Associations between neuropsychiatric/cognition measures and PDAQ scores stratified by sex**. Scatterplots illustrate the relationships between z-scored neuropsychiatric variables (GDS, BIS, AES, SHAPS, QUIP), cognitive variables (ExecFxn, Attn/WM)) and PDAQ score, stratified by sex **(A)** interaction analysis in the discovery sample **(B)** interaction analysis in the replication sample. Beta estimates and p-values represent the interaction of Sex*Neuropsychiatric/Cognition on PDAQ. Linear regression lines with 95% confidence intervals are overlaid.

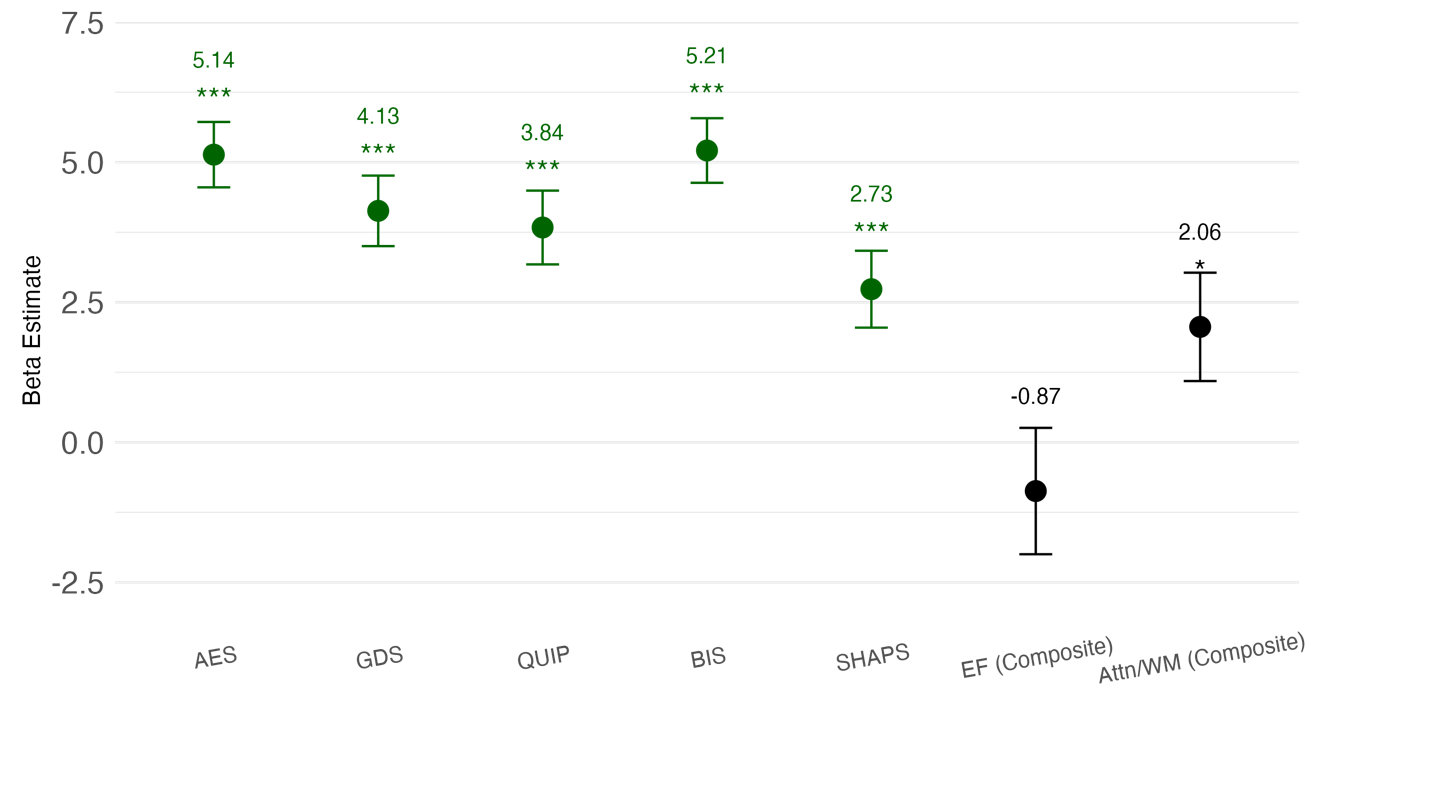

**Supplementary Figure 4**. **Standardized beta estimates of the association of neuropsychiatric and cognitive scores with PDAQ in the replication sample**. Regression coefficients (beta estimates) and 95% confidence intervals are shown for each predictor and are each derived from separate linear regression models that controlled for sex. Each predictor was z-scored prior to analysis, allowing effect sizes to be interpreted as the change in PDAQ score associated with a 1 SD increase in each predictor. Predictors are color-coded by domain: mood-related (dark green) and cognitive (black). Abbreviations: Executive Function (EF), Attention/Working Memory (Attn/WM).

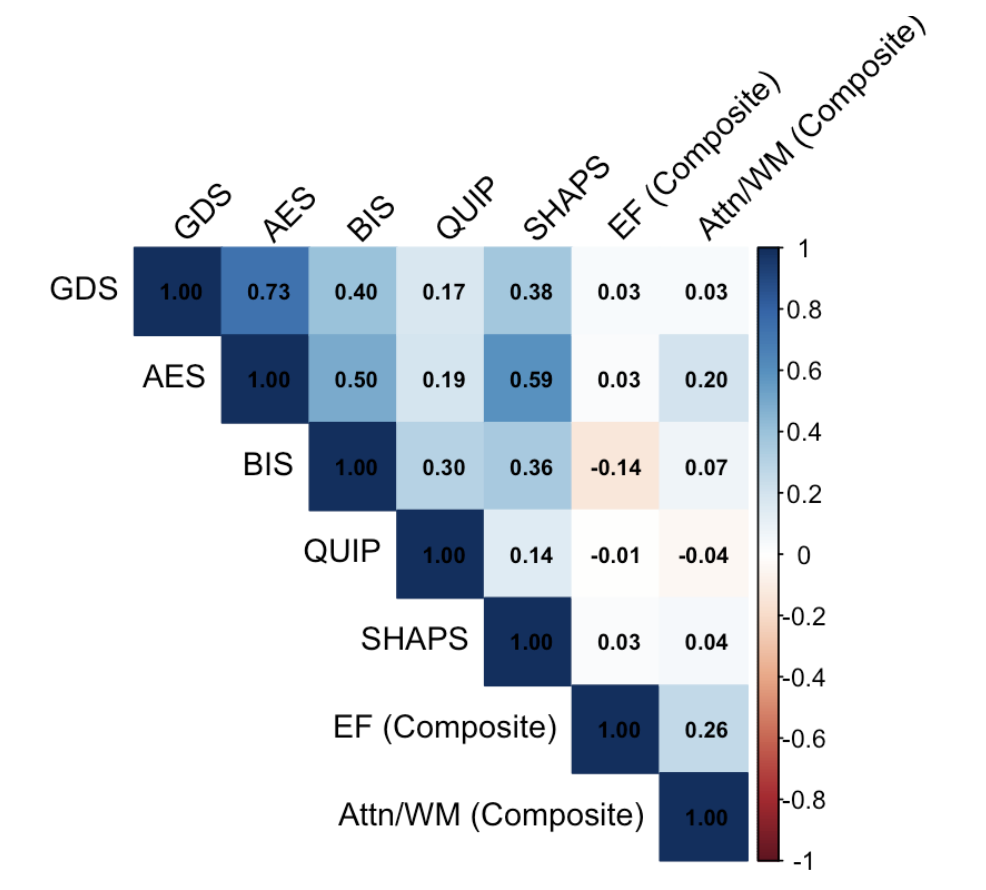

**Supplementary Figure 5.** **Correlation matrix of neuropsychiatric and cognition predictors in the replication sample**. Pairwise Pearson coefficients are displayed for all mood and cognition predictors included in the regression analyses. Coefficients are color-coded by strength and direction of association with darker blue indicating stronger positive correlations and red indicating negative correlations. Cognitive variables (EF, Attn/WM) show weak to no correlation with mood-related variables (AES, GDS, BIS, QUIP, SHAPS), indicating low collinearity across mood and cognition.

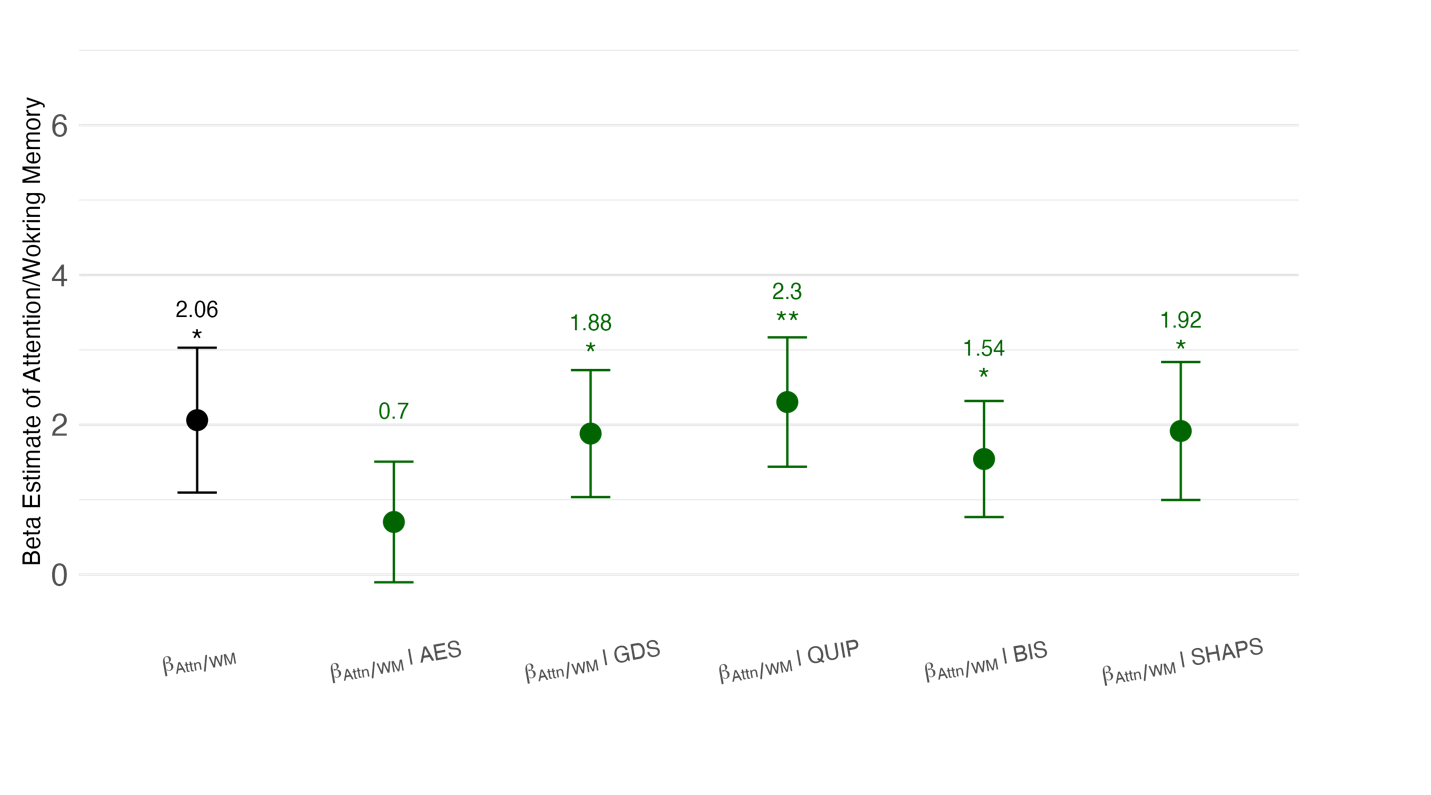

**Supplementary Figure 6. Assessment of confounding through comparison of the effect sizes for the association between cognition and PDAQ depending on the addition of neuropsychiatric scores in the replication sample**. Beta estimates for the effect of attention/working memory (Attn/WM) on PDAQ while controlling for mood predictors. In both models, all variables were z-scored to allow standardized effect size comparisons. Error bars represent 95% confidence intervals

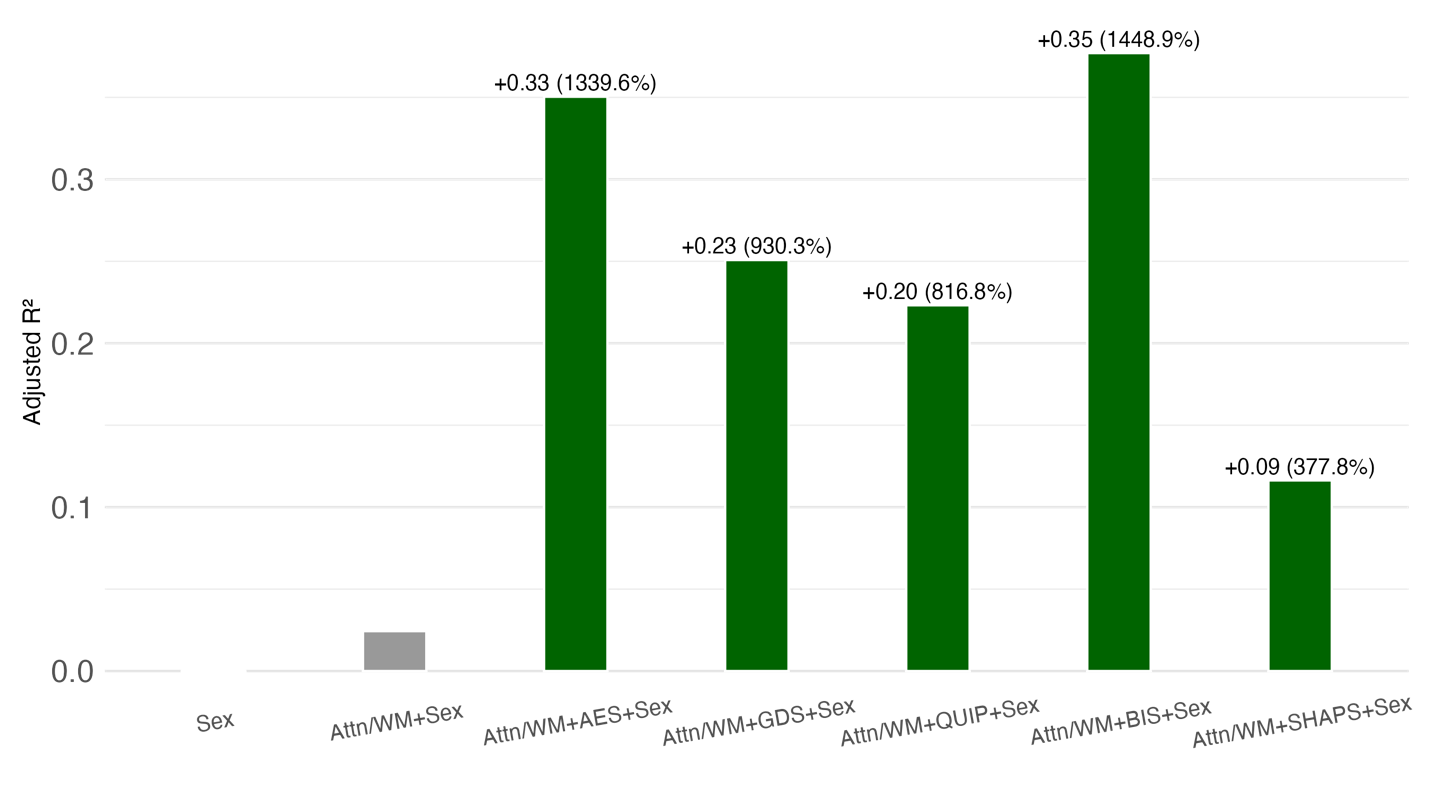

**Supplementary Figure 7. Comparison of proportion of variance explained in PDAQ by additive models using sex, cognition, and mood as predictors in the replication sample**. Models including sex alone, then sex and attention/working memory (Attn/WM), and then with the addition of each individual mood variable. Each bar represents the adjusted R² of the linear regression model. Text labels above each bar represent the absolute and relative increase in explained variance in models that add each of the mood measures compared to the base model that includes the cognitive measure and sex only.

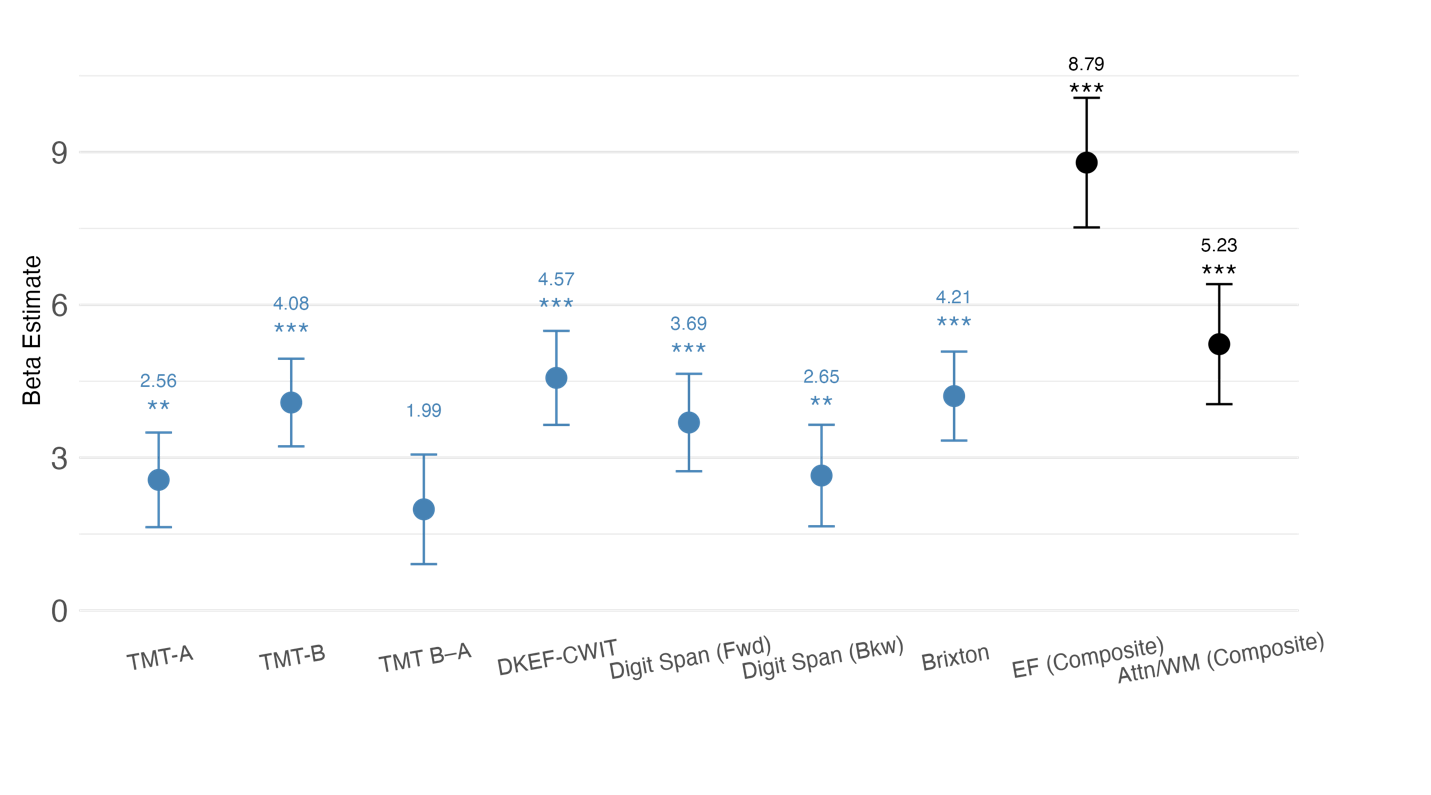

**Supplementary Figure 8**. **Standardized β estimates (95% CI) for the relationship between cognitive measures and PDAQ scores in the discovery sample**. Predictors include Trail Making Test (TMT-A, TMT-B, TMT B–A), D-KEFS Color-Word Interference Test (Stroop), Digit Span (Forward, Backward), and composite scores for Executive Function (EF) and Attention/Working Memory (A/WM). Predictors are color-coded by domain: individual cognitive measures (blue) and composite score (black).

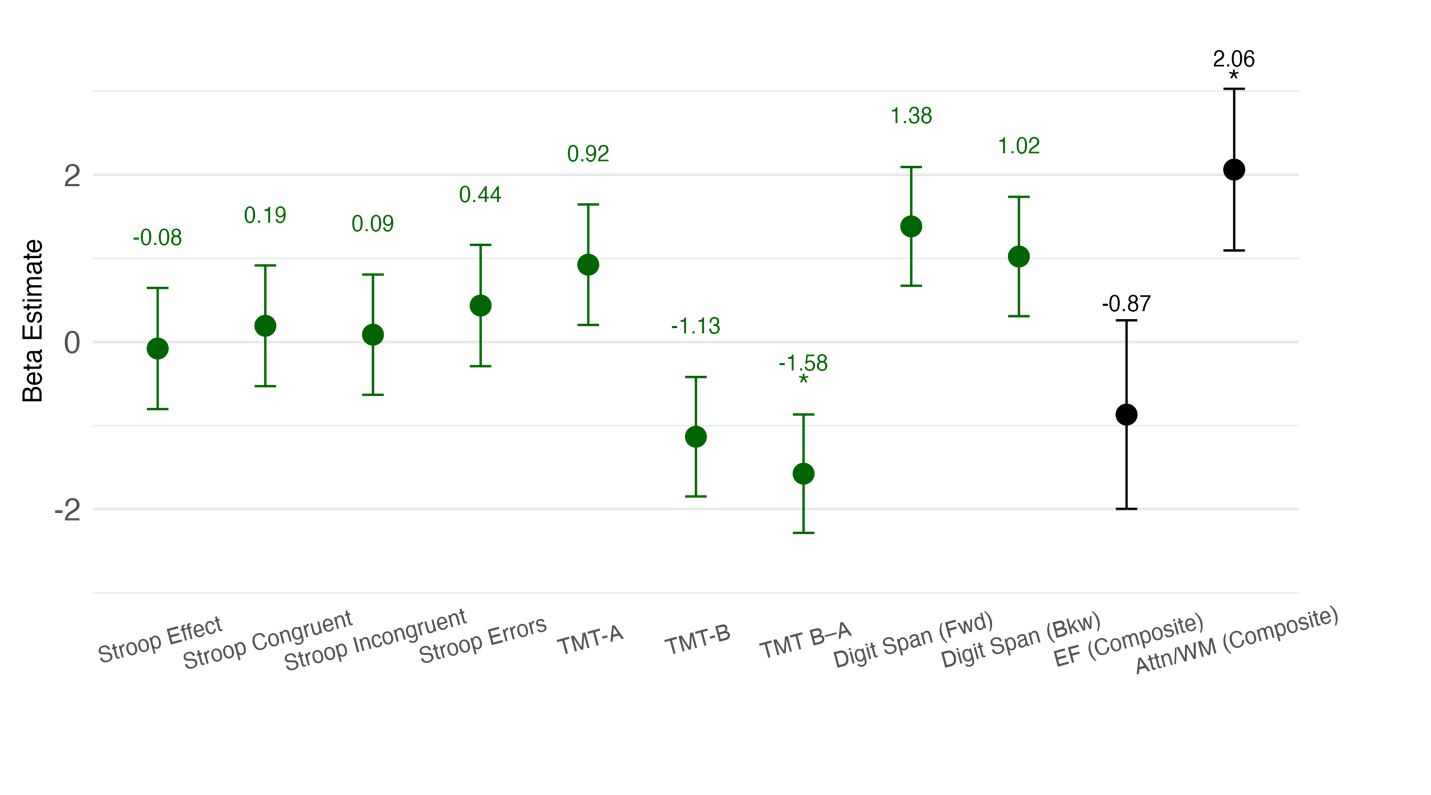

**Supplementary Figure 9**. **Standardized β estimates (95% CI) for the relationship between cognitive measures and PDAQ scores in the replication sample**. Predictors include Color-Word Stroop task (Stroop: Basic, Congruent, Incongruent, Errors), Trail Making Test (TMT-A, TMT-B, TMT B–A), Digit Span (Forward, Backward), and composite scores for Executive Function (EF) and Attention/Working Memory (A/WM). Predictors are color-coded by domain: cognitive measures (green) and composite score (black).
